# Exploring the Potential of ChatGPT-4 for Clinical Decision Support in Cardiac Electrophysiology and Its Semi-Automatic Evaluation Metrics

**DOI:** 10.1101/2024.07.10.24310247

**Authors:** Xiarepati Tieliwaerdi, Abulikemu Abuduweili, Saleh Saleh, Erasmus Mutabi, Michael Rosenberg, Emerson Liu

**Affiliations:** Department of Medicine, Allegheny Health Network; Robotics Institute, Carnegie Mellon University; Division of Cardiology, University of Colorado School of Medicine; Cardiovascular Institute, Allegheny Health Network

## Abstract

**Background/Aim:** Despite extensive research in other medical fields, the capabilities of ChatGPT-4 in clinical decision support within cardiac electrophysiology (EP) remain largely unexplored. This study aims to enhance ChatGPT-4’s domain-specific expertise by employing the Retrieval-Augmented Generation (RAG) approach, which integrates up-to-date, evidence-based knowledge into ChatGPT-4’s foundational database. Additionally, we plan to explore the use of commonly used automatic evaluation metrics in natural language processing, such as BERT score, BLEURT, and cosine similarity, alongside human evaluation, to develop a semi-automatic framework. This aims to reduce dependency on exhaustive human evaluations, addressing the need for efficient and scalable assessment tools in medical decision-making, given the rapid adoption of ChatGPT-4 by the public.

**Method:** We analyzed five atrial fibrillation (Afib) cases and seven cardiac implantable electronic device (CIED) infection cases curated from PubMed case reports. We conducted a total of 120 experiments for Afib and 168 for CIED cases, testing each case across four temperature settings (0, 0.5, 1, 1.2) and three seed settings (1, 2, 3). ChatGPT-4’s performance was assessed under two modes: the Retrieval-Augmented Generation (RAG) mode and the Cold Turkey mode, which queries ChatGPT without external knowledge via RAG. For Afib cases, ChatGPT was asked to determine rate, rhythm, and anticoagulation options, and provide reasoning for each. For CIED cases, ChatGPT is asked to determine the presence of device infections. Accuracy metrics evaluated the determination component, while reasoning was assessed by human evaluation, BERTScore, BLEURT, and cosine similarity. A mixed effects analysis was used to compare the performance under both models across varying seeds and temperatures. Spearman’s rank correlation was used to explore the relationship between automatic metrics and human evaluation.

**Results:** In this study, 120 experiments for Afib and 168 for CIED were conducted. There is no significant difference between the RAG mode and the Cold Turkey mode across various metrics including determination accuracy, reasoning similarity, and human evaluation scores, although RAG achieved higher cosine similarity scores in Afib cases (0.82 vs. 0.75) and better accuracy in CIED cases (0.70 vs. 0.66), though these differences were not statistically significant due to the small sample size. Our mixed effects analysis revealed no significant effects of temperature or method interactions, indicating stable performance across these variables. Moreover, while no individual evaluation metric, such as BERT score, BLEURT or cosine similarity, showed a high correlation with human evaluations. However, the ACC-Sim metric, which averages accuracy and cosine similarity, exhibits the highest correlation with human evaluation, with Spearman’s ρ at 0.86 and a P value < 0.001, indicating a significant ordinal correlation between ACC-Sim and human evaluation. This suggests its potential as a surrogate for human evaluation in similar medical scenarios.

**Conclusion:** Our study did not find a significant difference between the RAG and Cold Turkey methods in terms of ChatGPT-4’s clinical decision-making performance in Afib and CIED infection management. The ACC-Sim metric closely aligns with human evaluations in these specific medical contexts and shows promise for integration into a semi-automatic evaluation framework.

## 1 Introduction

Large language models (LLMs) are advanced AI systems designed to understand and generate human-like text based on vast amounts of data^1^. Among these, OpenAI’s ChatGPT-4 stands out as one of the most popular and widely recognized LLMs. In the medical field, ChatGPT-4 has achieved remarkable milestones, such as passing the United States Medical Licensing Examination (USMLE) and supporting general medicine by offering diagnostic suggestions, treatment options, and patient management advice^2^. Despite its success in general medicine, the application of ChatGPT-4 in specialized areas like electrophysiology (EP) remains largely unexplored. To enhance ChatGPT-4’s domain-specific knowledge, the Retrieval-Augmented Generation (RAG) approach can be employed^3^. RAG integrates external knowledge into the model, enabling it to address complex and niche queries with greater accuracy and relevance, transforming ChatGPT-4 from a general model into a specialized assistant^4,5^.

As ChatGPT’s use expands in high-stake fields like medicine, there is an urgent need for robust evaluation metrics to assess its performance accurately. Traditional evaluation methods, relying heavily on human involvement, can be time-consuming and burdensome, especially for experts facing burnout. To address this, metrics used for semantic contextual similarity evaluation such as BERT score, BLEURT, and cosine similarity have shown promise in machine translation and hold potential for assessing ChatGPT’s responses^6,7^. BERT score evaluates semantic similarity between generated and reference texts using contextual embeddings^8^. BLEURT, a BERT-based metric fine-tuned on human judgment data, enhances quality assessment accuracy^9^. Cosine similarity measures how closely related two pieces of text are, based on their vector representations^10^. These metrics provide scalable solutions for evaluating ChatGPT’s responses, reducing the reliance on exhaustive human evaluation.

This manuscript explores the clinical decision support potentials of ChatGPT-4 in EP, comparing the performance of using RAG to relying solely on its foundational database without providing external knowledge. Additionally, this study aims to explore the feasibility of employing automatic evaluation metrics commonly used in natural language processing to assess the reasoning of ChatGPT in the EP domain and its correlation with human evaluations. Our goal is to develop a semi-automatic framework that could alleviate the burden on human experts, particularly as the use of ChatGPT for medical decision-making expands, generating an increasingly large volume of information.

## 2 Method

### Design

This study employs ChatGPT to address EP-related medical queries, focusing on conditions such as atrial fibrillation (Afib) and cardiac implantable electronic device (CIED) infection. We chose these two topics because both Afib and CIED infections cover critical aspects of EP, including medical and procedural arrhythmia management and its complications, making them suitable for testing ChatGPT’s breadth of knowledge in EP. We aim to enhance the performance of ChatGPT in the EP domain using RAG. Additionally, we explore the development of a semi-automatic evaluation metric to assess the performance of ChatGPT addition to human scoring.

### Data and Case

In this study, we examined five cases related to Afib and seven cases related to CIED infection. They were curated from PubMed case reports^11–20^. For each Afib case, upon providing the specific case information, ChatGPT was tasked with offering recommendations on three critical aspects of care: determining rate control options, rhythm control options, and the necessity for and options regarding anticoagulation. Additionally, ChatGPT was required to provide reasoning for each decision it made. For each case, ‘determination’ involves selecting from provided choices, characterizing it as a single-choice problem, while ‘reasoning’ entails explaining about why it chose a particular option. One example of Afib cases is outlined in Table 1.

**Table 1.** Afib Questions.

| Afib - Questions |  | Answer Template |
| --- | --- | --- |
| Rate control options | Determination | Choices: "Beta blockers , calcium channel blockers" or "Amiodarone" , "Cardioversion" or "All of the above" or "None of the above" or "Rate control is not necessary" |
|  | Reasoning | Example: Cardioversion, because patient is hemodynamically unstable. |
| Rhythm control options | Determination | Choices: "Amiodarone" or "Catheter ablation" or "Cardioversion" or "All of the above" or "Rhythm control is not necessary" |
|  | Reasoning | Example: Rhythm control is not necessary, because patient is asymptomatic. |
| Anticoagulation necessity and options | Determination | Choices: "Unable to determine given available information" or "Anticoagulation is necessary with warfarin only" or "Anticoagulation is necessary with either warfarin or direct oral anticoagulants" or "Anticoagulation is not necessary" |
|  | Reasoning | Example: Unable to determine given available information, because information provided is not enough to calculate CHA<sub>2</sub>DS<sub>2</sub>-VASc score |

For each CIED case, after presenting the details, ChatGPT was tasked with addressing two critical aspects of care: determining whether the patient has a CIED infection, and deciding whether the CIED needs to be removed immediately or if removal can be deferred. Additionally, ChatGPT was required to provide reasoning for each decision made. For each aspect, ‘Determination’ involves selecting from provided choices, making it a single-choice problem, whereas ‘Reasoning’ involves providing the rationale behind the determination. One example of CIED cases is demonstrated in Table 2.

**Table 2.**
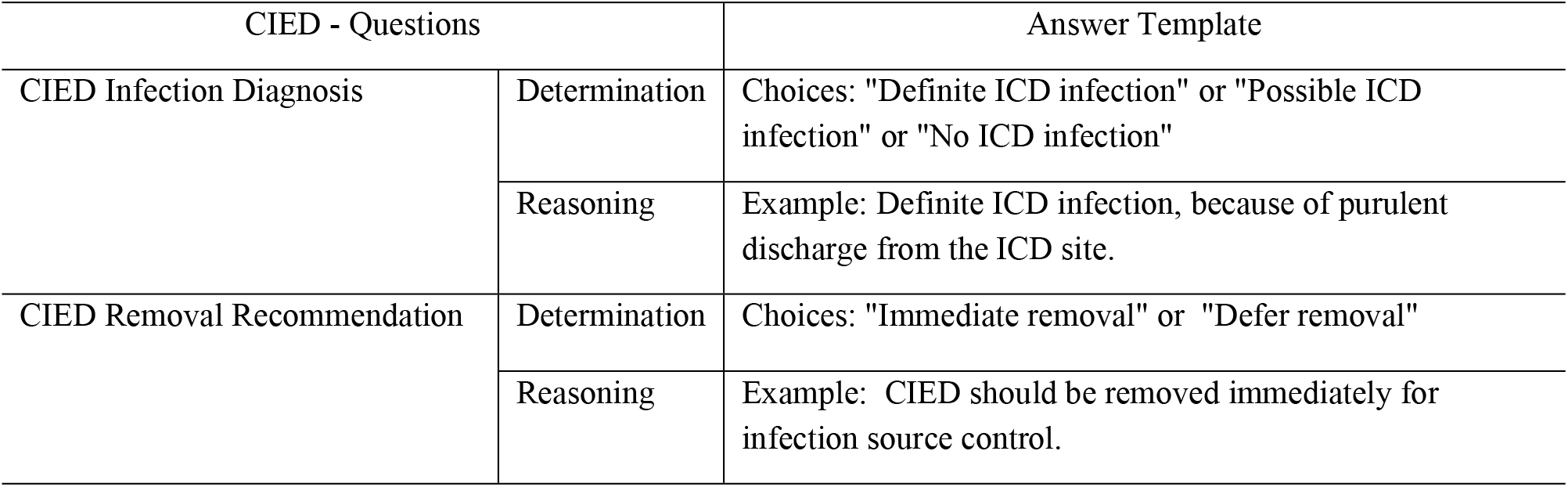
CIED Questions.

### Settings

In our study, we use “gpt-4-turbo-preview” version of ChatGPT. For each case, we conducted experiments using four different ‘temperature’ settings for ChatGPT. The ‘temperature’ parameter balances ChatGPT’s exploratory and conservative response capabilities, directly influencing the randomness of its responses. A higher temperature results in more random and creative responses, while a lower temperature leads to more deterministic outputs^21^. For each temperature setting, experiments were also performed using three different random seeds. A random seed is an initial value used by a random number generator to start the sequence of random numbers. Using the same seed will generate the same sequence of random numbers, which will decrease the randomness of the model and enhance consistency when the same tasks are repeated multiple times^22^. Consequently, for each case, we conducted a total of 12 trials per method (4 temperature settings *×* 3 seeds).

### RAG

In this study, we implemented Retrieval-Augmented Generation (RAG) as our foundational prompting technique. RAG optimizes the output of a large language model by referencing an external authoritative knowledge base beyond its initial training data before generating a response^3^. We constructed a knowledge library containing 10 guidelines for CIED from AHA^23^ and UptoDate ^24^ and 50 guidelines for Afib from ACC/AHA/ACCP/HRS^25^ and UpToDate^26,27^. Using RAG, we identified the top-5 most relevant guidelines from the library by calculating the similarity between the guidelines and the current case. Specifically, we utilized OpenAI’s embedding model ‘text-embedding-3-small’ to generate vector embeddings for both the guidelines and case descriptions. We then applied cosine similarity to these vectors to determine the most pertinent guidelines based on the highest similarity scores.

For comparison, we also conducted experiments using the baseline ChatGPT-4 querying method, termed the ‘Cold Turkey’ approach, which involves directly querying ChatGPT-4 without the aid of RAG or the knowledge library.

### Evaluation metrics

We evaluated the performance of the ‘determination’ problem, which consists of single-choice questions, using accuracy metrics. For ‘reasoning,’ we employed an embedding-based similarity score to evaluate the rationale. Specifically, we utilized the OpenAI text embedding model, version “ `text-embedding-3-small”` , to convert both the standard reasoning from human expert and the ChatGPT-provided reasoning into 1536-dimensional vectors. We then measured the similarity between these two vectors using cosine similarity to evaluate the correspondence between the actual reasoning and the response provided by ChatGPT. For a more rigorous evaluation, we also implemented human evaluation to assess the accuracy of ChatGPT’s responses. In this evaluation, an internal medicine resident rates ChatGPT’s responses based on the following criteria: a score of 0 for incorrect answers, 0.5 for correct determination but incorrect reasoning, 0.75 for correct determination and reasonable but incomplete reasoning, and 1.0 for both correct determination and fully accurate reasoning.

### Automatic Evaluation

To explore automatic evaluation methods in addition to human experts, we investigated the correlation between several automatic metrics and human evaluation scores. In addition to the previously discussed determination accuracy and cosine similarity, we included the BERTScore and BLEURT Score, which are widely used metrics for assessing the semantic similarity between two texts. These metrics utilize embeddings from BERT language model to measure the alignment between texts^8,9^.

### Statistical Analysis

Linear mixed-effects models were used to evaluate the effect of different prompting techniques (RAG and Cold Turkey) and to assess the impact of temperature and random effects. In this study, we utilized the Python package `statsmodels` (version 0.14) to conduct the linear mixed effects analysis^28^. `SciPy` (version 1.14) was used to conduct Spearman’s rank correlation^29^.

## 3 Results

We analyzed five Afib cases and seven cases concerning CIED infections. These cases were tested across four different temperature settings and three different seeds. This experimental setup was applied under both Cold Turkey mode and RAG mode. As a result, a total of 120 experiments were conducted for Afib cases and 168 experiments for CIED cases. Details of the scoring records can be found in the supplementary materials.

### Performance of RAG

Table 3 presents a performance comparison between the Cold Turkey and RAG methods for Afib cases, evaluating determination accuracy, cosine similarity, and human evaluation scores. The results are reported in the format of mean ± standard deviation. As illustrated, RAG consistently outperforms Cold Turkey across all metrics on average, however it didn’t reach statistical significance. Table 4 illustrates the performance comparison between the Cold Turkey and RAG methods for CIED cases. Although there is a trend of RAG consistently outperforming Cold Turkey across all metrics on average, these differences did not reach statistical significance.

**Table 3.** Performance Comparison of Cold Turkey and RAG methods for Afib cases.

| Metrics | Determination Accuracy |  | Cosine Similarity |  | Human Evaluation Score |  |
| --- | --- | --- | --- | --- | --- | --- |
|  | Cold Turkey | RAG | Cold Turkey | RAG | Cold Turkey | RAG |
| Rate Control Options | $0.39 \pm 0.49$ | $0.38 \pm 0.41$ | $0.74 \pm 0.1$ | $0.79 \pm 0.09$ | $0.34 \pm 0.43$ | $0.34 \pm 0.40$ |
| Rhythm Control Options | $0.66 \pm 0.47$ | $0.66 \pm 0.41$ | $0.74 \pm 0.08$ | $0.82 \pm 0.08$ | $0.50 \pm 0.41$ | $0.56 \pm 0.45$ |
| Anticoagulation Necessity Options | $0.50 \pm 0.50$ | $0.59 \pm 0.46$ | $0.78 \pm 0.09$ | $0.85 \pm 0.1$ | $0.44 \pm 0.46$ | $0.51 \pm 0.47$ |
| Average | $0.52 \pm 0.11$ | **$0.54 \pm 0.13$** | $0.75 \pm 0.02$ | **$0.82 \pm 0.03$** | $0.42 \pm 0.07$ | **$0.47 \pm 0.11$** |

**Table 4.**
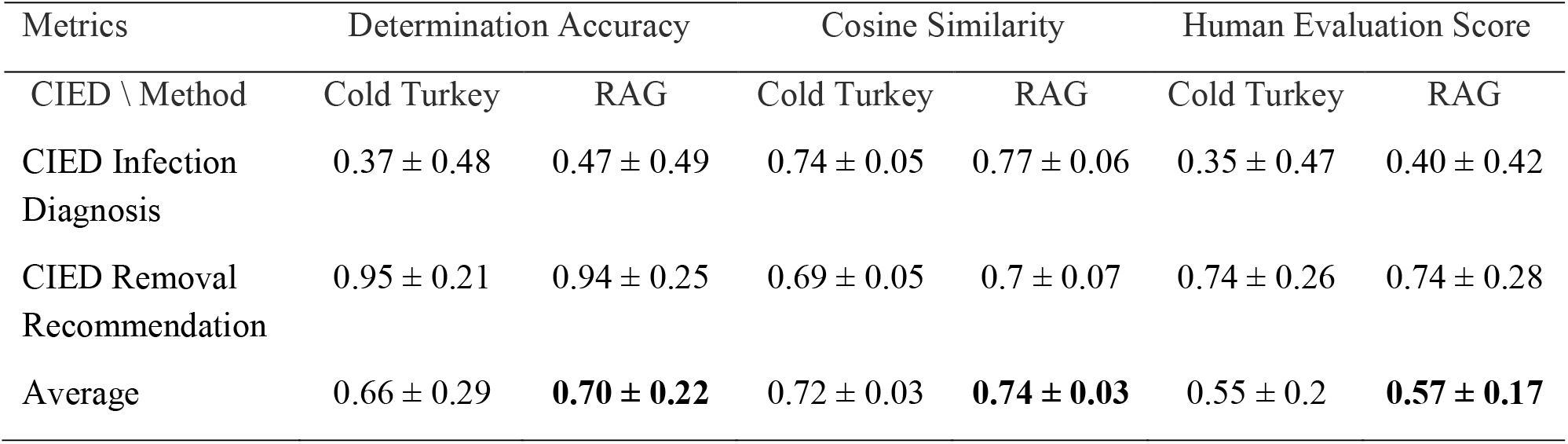
Performance Comparison of Cold Turkey and RAG methods for CIED cases.

### Automatic Evaluation

We measured Spearman’s rank correlation between various automatic metrics and human evaluation scores. The automatic metrics included accuracy of determination, cosine similarity, BERT score, BLEURT, and ACC-Sim, where ACC-Sim represents the average of accuracy and cosine similarity. Table 6 reports the 5 Spearman’s ρ and the corresponding p-value in parentheses between several metrics and human evaluation scores. As shown, ACC-Sim exhibits the highest correlation with human evaluation scores, with a P value < 0.001, indicating a significant ordinal correlation between ACC-Sim and human evaluation.

**Table 6.**
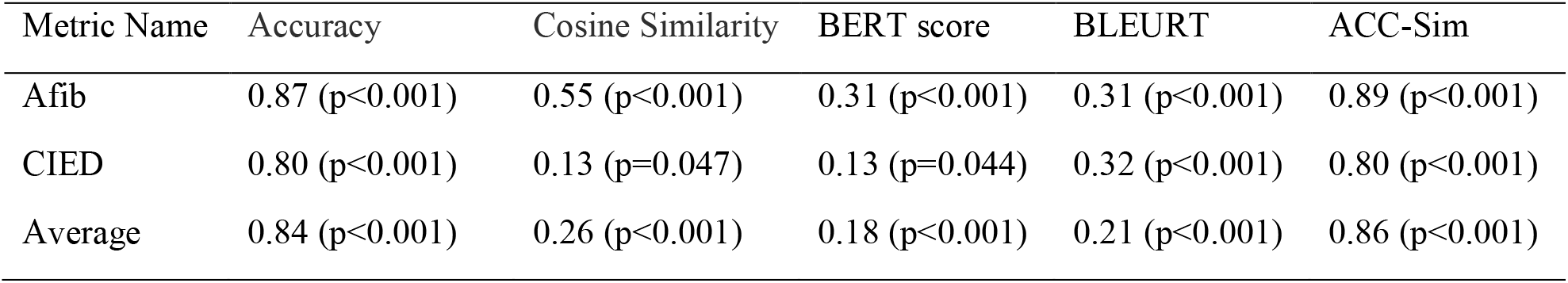
Spearman’s *ρ* between several automatic metrics and human evaluation scores.

We illustrate the relationship between ACC-Sim and human evaluation scores in Figure 1. The yellow points represent the ACC-Sim and human evaluation scores from different experimental trials. The black curve represents the ordinal logistic regression fit of the relationship, using ACC-Sim as the input and human evaluation as the output. ACC-Sim effectively captures the ordinal nature of human evaluation: higher ACC-Sim scores correspond to higher scores, and vice versa.

**Figure 1.**
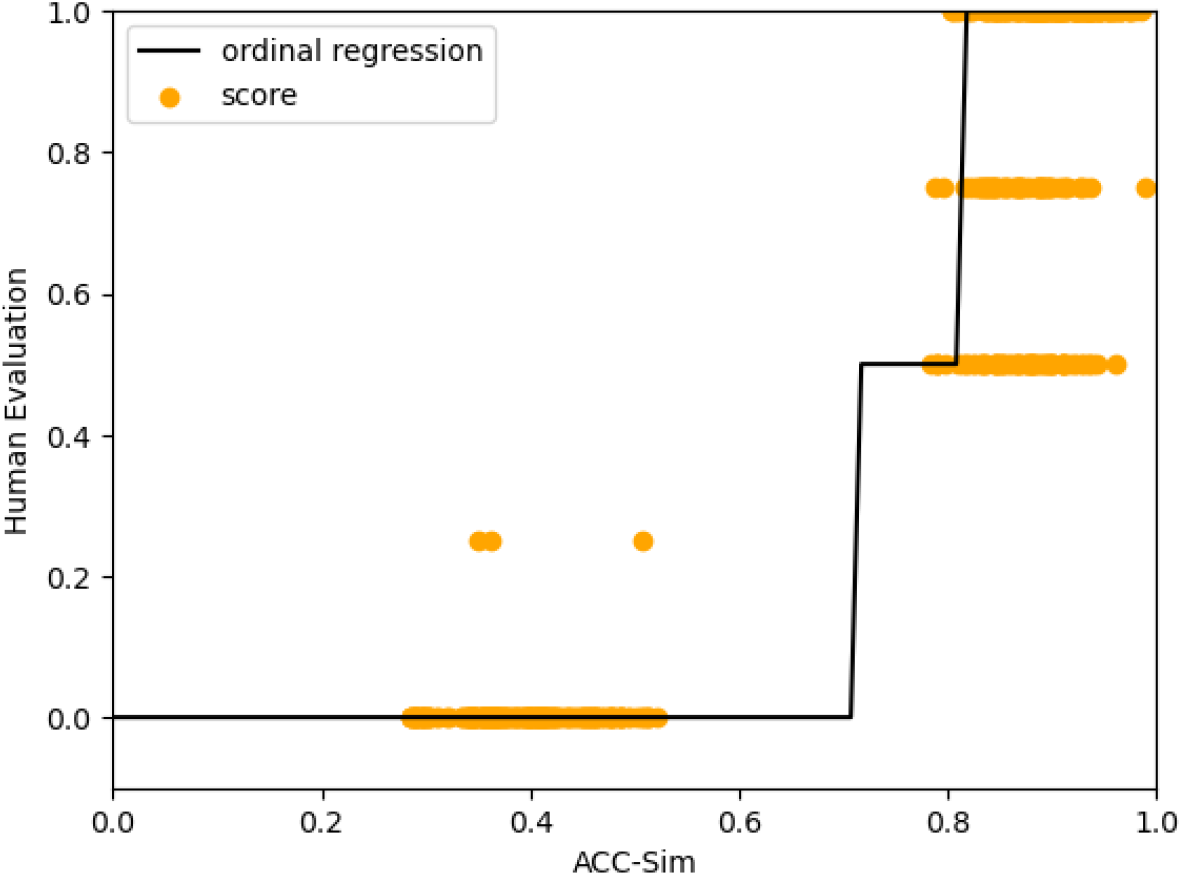
Relationship between ACC-Sim and Human Evaluation scores.

### Mixed Effect Analysis

Table 3 and Table 4 demonstrate that there is a trend that RAG generally outperforms Cold Turkey on average. In this section, we employed a Mixed Effects Model to analyze the impact of various factors. We defined the prompting technique (RAG or Cold Turkey) and temperature as fixed effects, while treating the random seed as a random effect to assess their relationships with the human evaluation score. Table 7 reports the Mixed Effect Analysis on Afib cases. As indicated in Table 7, the Intercept (Coefficient=0.454) reflects the baseline effect when using the ‘Cold Turkey’ method at zero temperature. The coefficient for the Technique [RAG] quantifies the difference in the response variable when employing the ‘RAG’ method compared to the ‘Cold Turkey’ approach. Specifically, Technique [RAG] (0.031, p=0.656) suggests that the RAG approach may achieve a slightly higher human evaluation score (+0.031) than the baseline; however, this difference is not statistically significant (p=0.656). On the other hand, as indicated by the right column, the 95% confidence interval for the Technique [RAG] coefficient estimate ranges from [-0.105, 0.167]. This interval is not strictly positive, making it difficult to assert that the RAG method is significantly superior to the Cold Turkey approach. Furtherly, the effect of temperature (-0.058, p = 0.496) is not statistically significant, indicating that temperature changes do not significantly impact the response variable. The group variance being close to zero indicates minimal variability in the response variable attributable to differences between seeds.

**Table 7.** Mixed Effect Analysis Results on Human Evaluation Score of Afib Case.

| Variable | Coefficient | Standard Error | P-value | [0.025 | 0.975] |
| --- | --- | --- | --- | --- | --- |
| Intercept [Cold Turkey] | 0.454 | 0.066 | 0.000 | 0.325 | 0.583 |
| Technique [RAG] | 0.031 | 0.069 | 0.656 | -0.105 | 0.167 |
| Temperature | -0.058 | 0.085 | 0.496 | -0.225 | 0.109 |
| Group Variance | 0.000 |  |  |  |  |

Table 8 reports the mixed effect analysis on CIED cases. Similar to the results of Afib, Technique [RAG] (0.025, p=0.612) suggests that the RAG approach may achieve a slightly higher human evaluation score than the baseline; however, this difference is not statistically significant (p=0. 612). Furtherly, the effect of temperature (0.035, p = 0.555) is not statistically significant, indicating that temperature changes do not significantly impact the outcome.

**Table 8.** Mixed Effect Analysis Results on Human Evaluation Score of CIED Case.

| Variable | Coefficient | Standard Error | P-value | [0.025 | 0.975] |
| --- | --- | --- | --- | --- | --- |
| Intercept [Cold Turkey] | 0.528 | 0.049 | 0.000 | 0.431 | 0.625 |
| Technique [RAG] | 0.025 | 0.049 | 0.612 | -0.071 | 0.120 |
| Temperature | 0.035 | 0.060 | 0.555 | -0.082 | 0.152 |
| Group Variance | 0.001 | 0.011 |  |  |  |

## 4 Discussion

This study evaluated ChatGPT-4’s performance in EP by testing its decision-making capabilities under both the RAG and the Cold Turkey mode. Our analysis involved five cases of Afib and seven cases of CIED infection, derived from PubMed case reports. These cases were thoroughly assessed across various conditions—four different temperature settings and three different seeds— resulting in a comprehensive dataset of 120 experiments for Afib and 168 for CIED.

Our study did not find a significant difference between the RAG and Cold Turkey methods. However, since the average score for the RAG method was slightly higher than that for the Cold Turkey method, increasing the sample size might reveal significant differences between these two methods. This is part of our ongoing work. The investigation into temperature effects showed no significant impact on the response variable, suggesting that ChatGPT responses are stable across different temperature settings.

In this study, determination accuracy showed a higher correlation with human evaluation compared to cosine similarity, BERT score, and BLEURT score when evaluating ChatGPT’s responses. By averaging determination accuracy and cosine similarity, we obtained the ACC-Sim metric, which is more closely correlated with human evaluation. ACC-Sim can effectively evaluate the performance of ChatGPT-4’ responses in Afib or CIED tasks and holds potential as a surrogate for human evaluation in these specific cases.

## 5 Conclusion

Our study did not find a significant difference between the RAG and Cold Turkey methods in terms of ChatGPT-4’s clinical decision-making performance in Afib and CIED infection management, although the average score for the RAG method was slightly higher than that for the Cold Turkey method. The ACC-Sim metric, which averages accuracy and cosine similarity, is highly correlated with human evaluations in these specific medical contexts and shows promise for integration into a semi-automatic evaluation framework. However, larger datasets are required to validate the comparative analyses between different prompting techniques and the correlation between automated scores and human ratings across a broader spectrum of clinical conditions. These automated metrics are far from being substitutes for human expertise.

## Data Availability

All data produced in the present study are available upon reasonable request to the authors.

## Notes

### Competing Interest Statement

The authors have declared no competing interest.

### Funding Statement

This study did not receive any funding.

### Author Declarations

The study used (or will use) ONLY openly available human data that were originally located at Pubmed case reports. https://pubmed.ncbi.nlm.nih.gov/

### Summary of Updates

Updates of corresponding authors' information and correction of erroneous citations.

